# Deep Learning Prediction of Personalized Peripapillary Retinal Nerve Fiber Layer Thickness Norms from Fundus Images in Glaucoma

**DOI:** 10.64898/2026.05.26.26354081

**Authors:** Elif Yildiz, Lucy Zha, Nazlee Zebardast, Min Shi, Mengyu Wang

## Abstract

**Purpose:** To predict retinal nerve fiber layer thickness (RNFLT) norms from fundus images.

**Methods:** We selected 18,000 OCT scans and visual fields (VF) from the Massachusetts Eye and Ear Glaucoma Service. A U-Net-based deep learning model was developed to predict RNFLT norms from OCT en face fundus images. A total of 10,000 OCT scans with normal VFs (mean deviation [MD] ≥ -1 dB, glaucoma hemifield test within normal limits, and pattern standard deviation probability > 5%) tested within 30 days were used for training, while the remaining 8,000 OCT scans (mean VF MD: −3.3 ± 4.9 dB), including 2,419 scans with normal VFs, were used for evaluation. Structure-function correlations between RNFLT maps and VFs were assessed using linear regression and VGG-16 across original RNFLT maps, deviation maps, and their combination. Performance was evaluated using correlation coefficients, mean absolute error (MAE), and R^2^.

**Results:** Predicted RNFLT norm maps showed agreement with baseline RNFLT maps in eyes with normal VFs (R^2^ = 0.81 ± 0.13). RNFLT deviation maps correlated more strongly with VF MD than original RNFLT maps (R = 0.42 vs. 0.19, p < 0.01). In deep learning-based VF prediction, combining original and deviation maps achieved the best performance (MAE = 3.31 dB, R^2^ = 0.39), outperforming the model (p < 0.05) using original RNFLT maps alone (MAE = 3.36 dB, R^2^ = 0.35).

**Conclusions:** Deep learning can estimate individualized RNFLT norms and improve structure-function assessment in glaucoma.

**Translational Relevance:** Personalized RNFLT norm prediction may improve detection of glaucomatous damage.

## Introduction

Glaucoma is a chronic eye disease characterized by progressive optic neuropathy that leads to irreversible vision loss.^1^ It is the second most common cause of blindness worldwide.^2^ Structurally, glaucoma is characterized by a gradual loss of retinal ganglion cells, resulting in thinning of the retinal nerve fiber layer (RNFL), while functionally, it causes irreversible visual field (VF) loss. Structural changes in the RNFL often precede functional loss, making retinal nerve fiber layer thickness (RNFLT) a crucial biomarker for early diagnosis and monitoring of glaucoma. ^3, 4, 5^ In routine clinical practice, retinal nerve fiber layer thickness (RNFLT) measurements are widely utilized by ophthalmologists for both the diagnosis and monitoring of glaucoma progression. To facilitate earlier recognition of structural alterations, clinicians often rely on normative deviation maps generated by comparing the patient’s RNFLT values to population-derived reference data. ^3^ These reference datasets are constructed from population-level data, stratified by age, sex, and sometimes ethnicity. Such normative datasets are used in OCT devices like Spectralis and Cirrus. However, they are generally built upon only basic demographic variables such as age and sex.^5,6^ It is increasingly recognized that individual anatomical variations of the retina significantly influence RNFLT and must be accounted for to improve diagnostic accuracy.

There are multiple retinal anatomical factors that affect the thickness of the retinal nerve fiber layer. One key factor is axial length variation associated with refractive errors. Myopic eyes, which tend to have longer axial lengths, often show thinner measurements of the RNFLT, potentially leading to false-positive diagnoses of glaucoma. In contrast, hyperopic eyes may appear to have thicker RNFLT values due to their shorter axial lengths, which can mask existing glaucoma damage and delay diagnosis. ^4, 7, 8^ Moreover, in healthy individuals, RNFL peak thickness locations are closely associated with the position of major retinal blood vessels, which can vary considerably between individuals. ^9, 10^ Other anatomical features such as disc-fovea angle ^11, 12^, optic disc torsion^13^, and disc tilt^14,15^ also play an important role in shaping normal RNFL profiles. More accurate personalized RNFLT norms are essential for improving diagnostic accuracy and disease monitoring. It is very important to adjust these RNFLT norms according to variable retinal anatomical parameters. In previous studies,^9,10,13^ retinal features such as vessel positioning and disc parameters were manually assessed for the evaluation of RNFL thickness. This was not preferred in practical use because it is time consuming, labor intensive, and subject to interobserver variability. Furthermore, although the impact of race on RNFL thickness norms has been studied extensively, race-specific personalized normative models have not been developed yet.

The aim of this study is to leverage deep learning models to predict patient-specific, expected normal RNFLT maps directly from fundus images captured via OCT. Because major fundus-visible anatomy, particularly the retinal vascular pattern, is highly stable over time, while glaucoma progression primarily manifests as RNFL thinning, models can leverage static fundus information to infer RNFLT profiles, consistent with prior work predicting OCT-derived RNFLT from fundus photographs.^4,16^ In other words, the developed model creates personalized RNFLT norms and compares them with the current RNFLT to provide a more accurate diagnosis and follow-up in glaucoma management with a personalized interpretation for the individual.

## Methods

### Data Description

This retrospective study was approved by the Mass General Brigham Institutional Review Board and determined to be exempt from the informed consent requirement. The research adhered to the tenets of the Declaration of Helsinki.

The OCT dataset was from the Massachusetts Eye and Ear glaucoma service between 2010 and 2022 (**Table 1**). This study included 18,000 reliable OCT scans (signal strength ≥ 6) from 17,835 eyes of 13,821 patients, with a mean age of 57.2 ± 16.1 years. Each RNFLT map consisted of 200 × 200 thickness values and was obtained from a spectral-domain OCT device (Cirrus, Carl Zeiss Meditec, Dublin, California). VFs were tested within 30 days of corresponding OCT scans acquisition, and VF reliability was defined as fixation loss ≤ 33%, false positive rate ≤ 20%, and false negative rate ≤ 20%. The mean deviation (MD) of 18,000 VFs was -1.4 ± 3.67 dB. The deep learning model was trained using 10,000 baseline OCT scans of eyes with VFs (MD: 0.06 ± 0.72 dB) tested within 30 days that were clinically normal meeting the criteria of MD ≥ -1 dB, glaucoma hemifield test within normal limits and pattern standard deviation probability > 5%. The remaining 8,000 OCT scans with a mean MD of -3.3 ± 4.9 dB (including 2,419 OCT scans with normal VFs, MD: 0.04 ± 0.73 dB) were used for evaluating the prediction performance.

**Table 1.** Dataset statistics.

| Characteristic | N= 13,821 patients |
| --- | --- |
| Number of patients (eyes) | 13,821 (17,835) |
| Number of OCT and VF pairs | 18,000 |
| Age (years) | $57.2 \pm 16.1$ |
| Percentage of right eyes | 53% |
| Average RNFLT | $88.24 \pm 19.11$ ($\mu$) |
| Average HVF MD | $-1.4 \pm 3.67$ dB |
| Number of training images | 10,000 (MD: $0.06 \pm 0.72$ dB) |
| Number of testing images | 8,000 (MD: $-3.3 \pm 4.9$ dB) |

All data are presented as mean ± standard deviation, unless otherwise stated.

### Deep Learning Model for RNFLT Norm Prediction

We developed a deep learning model using a UNet-based architecture to predict RNFL maps from en face fundus images obtained from OCT scans (**Figure 1**). The RNFLT norm or deviation map was obtained via three consecutive steps: (1) first, we train a UNet-based autoencoder model, which inputs en face fundus images and outputs corresponding normal RNFLT maps. Only OCT scans with normal VFs were used for model training; (2) Then, the pretrained model is used to predict corresponding normal RNFLT maps from OCT fundus images of eyes selected without restricting their VF MD values; (3) Finally, the RNFLT deviation map was calculated as the difference between predicted normal RNFLT map and RNFLT map matched with the en face fundus image (i.e. the model input to predict normal RNFLT map). We evaluated the prediction performance by correlating the predicted RNFLT map and corresponding ground-truth normal (baseline) RNFLT map.

**Figure 1.**
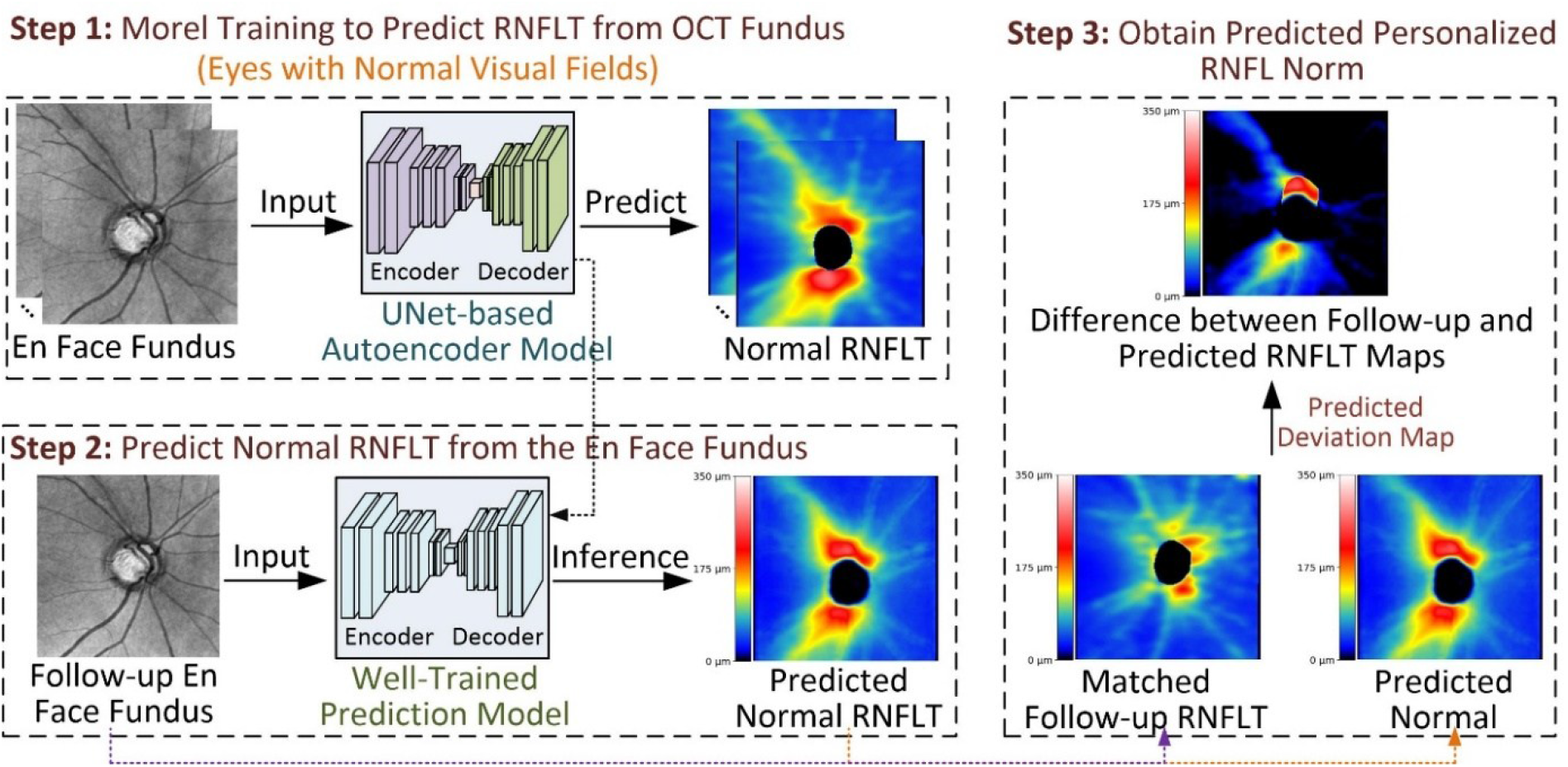
Proposed framework for predicting personalized RNFLT normative maps from en face OCT fundus images and deriving RNFLT deviation maps.

### Structure-Function Correlation Assessment

We linearly correlated the mean thickness value of the RNFLT deviation map with the corresponding VF MD and compared it against the correlation achieved by the original RNFLT map. In addition, we tested if the predicted RNFLT maps have better correlation with VFs quantified by deep learning models (VGG-16) compared with original RNFLT maps by three sets of experiments: (1) 52 VF total deviation (TD) values prediction from original RNFLT maps; (2) 52 VF TDs prediction from RNFLT deviation maps; and (3) 52 VF TDs prediction from the combination of original RNFLT map and RNFLT deviation map. Mean absolute error (MAE) and R-squared (R^2^) were used to measure the structure-function correlation.

### Data Analysis and Statistics

We used the t-test combined with three-fold cross-validation. For the three-fold cross-validation, the data were split into three equal subsets, with each subset used iteratively for model evaluation and the remaining two subsets combined for model training. The bootstrapping technique was used to compare the structure-function correlation between original RNFLT maps and predicted RNFLT maps. All statistical analyses were performed using Python 3.8.

## Results

### RNFLT Norm Prediction Results

On the 2,419 RNFLT with normal VFs, when comparing baseline RNFLT maps and predicted RNFLT maps, the model achieved an average correlation (R) of 0.91 and mean absolute error (MAE) of 12.75 microns (**Figure 2**). The R^2^ followed a tight distribution on all predictions with average as 0.81 ± 0.13 (**Figure 3**). For comparison, we additionally tested using Scanning Laser Ophthalmoscopy (SLO) fundus and Inner Limiting Membrane (ILM) maps to predict RNFL norm, respectively (**Figure 4**). SLO fundus images achieved an R correlation of 0.73 and MAE of 19.54 microns, while ILM maps yielded an R correlation of 0.9 and MAE of 16.44 microns.

**Figure 2.**
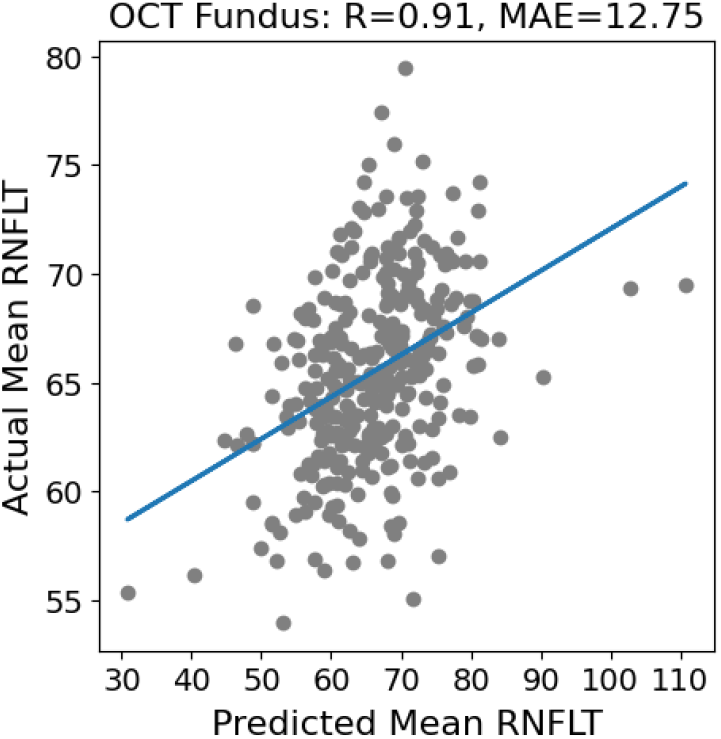
Agreement between baseline RNFLT maps and predicted normative RNFLT maps in eyes with normal visual fields.

**Figure 3.**
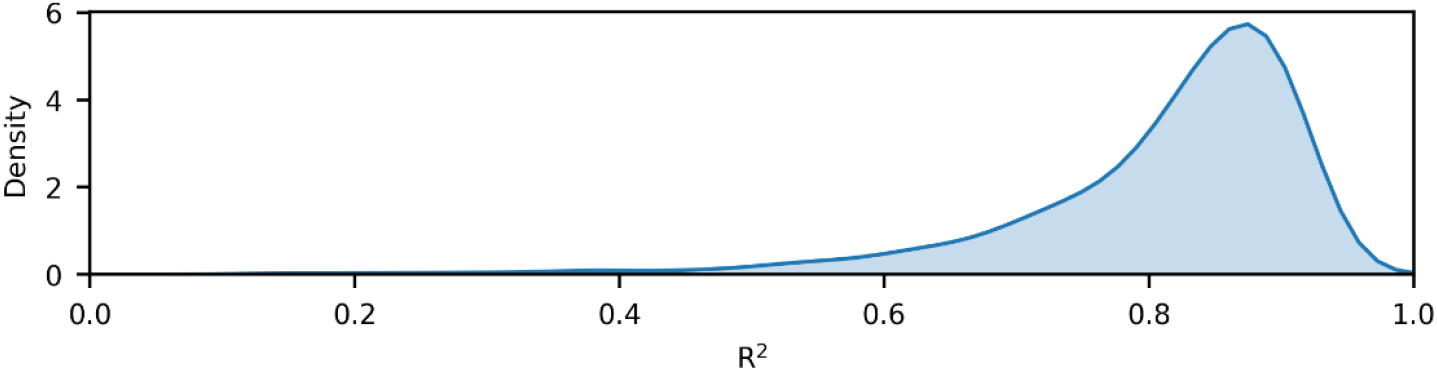
Distribution of R^2^ values for normative RNFLT map prediction in 2,419 OCT scans with normal visual fields.

**Figure 4.**
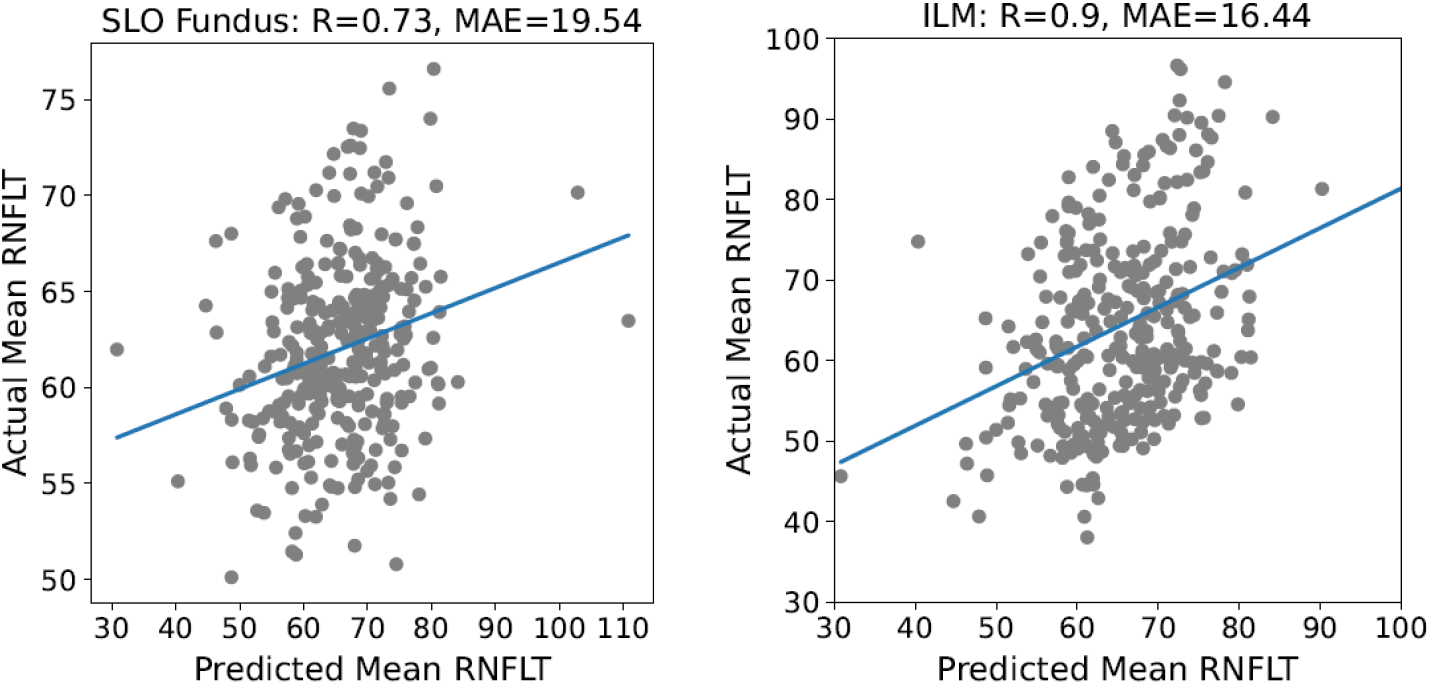
Correlation of original normal RNFLT maps and RNFLT maps predicted using SLO fundus and ILM maps to predict RNFL norm, respectively.

### Structure-Function Correlation Results

Based on the linear regression model, the correlation coefficient (R) of average RNFLT deviation values with VF MDs was 0.42, which almost doubled the correlation score (R = 0.19) achieved by the average original RNFLT values (p-value < 0.01). With the deep learning model, both MAE and R^2^ consistently demonstrate that combining original RNFLT maps with RNFLT deviation maps yields the strongest structure-function correlation for predicting 52 VF TD values (**Figure 5** and **Figure 6**). Using original RNFLT maps alone resulted in a mean MAE of 3.36 dB and a mean R^2^ of 0.35, while RNFLT deviation maps alone showed slightly inferior performance with a mean MAE of 3.40 dB and a mean R^2^ of 0.34. In contrast, the combined RNFLT representation achieved the lowest prediction error (mean MAE = 3.31 dB) and the highest explanatory power (mean R^2^ = 0.39) across most VF locations.

**Figure 5.**
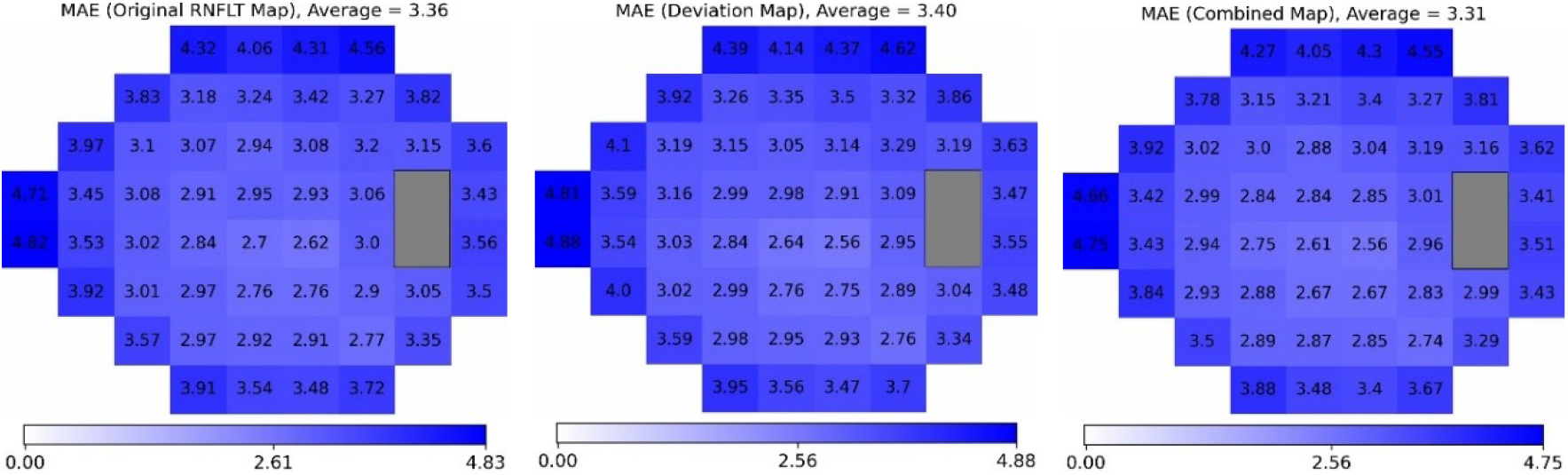
Mean Absolute Error (MAE) results of TDs prediction from original RNFLT map, RNFLT deviation map, and combined (original + deviation) RNFLT map.

**Figure 6.**
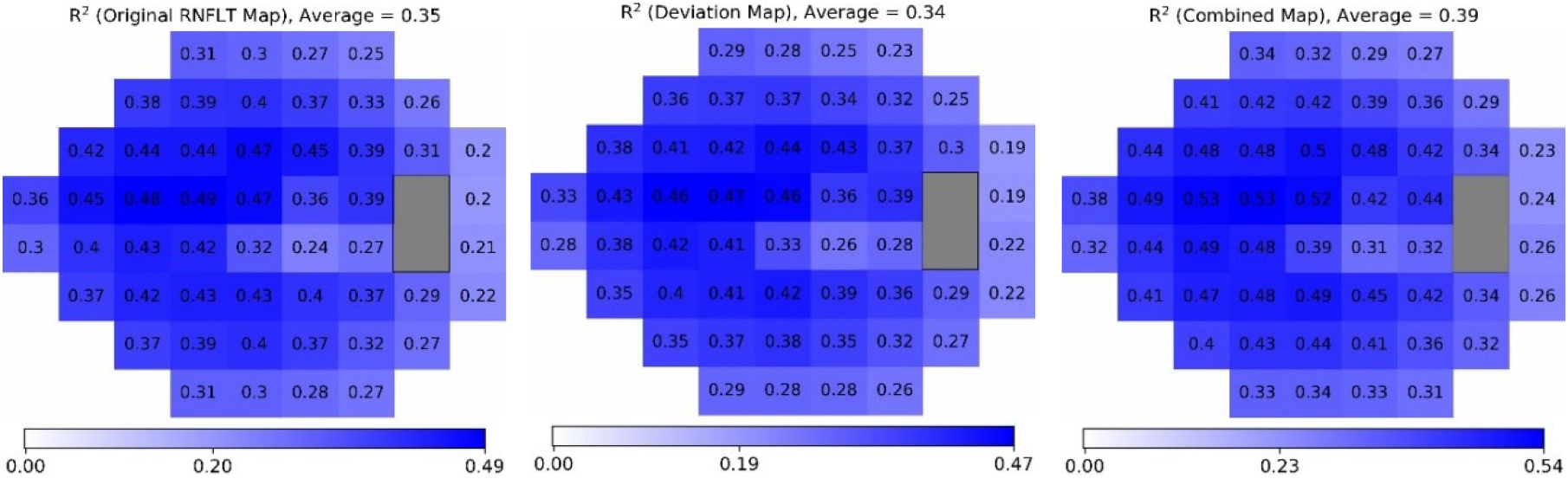
R^2^ results of TDs prediction from original RNFLT map, RNFLT deviation map, and combined (original + deviation) RNFLT map.

## Discussion

This study demonstrates the feasibility and clinical value of predicting personalized RNFLT norms from en face OCT fundus images using deep learning. By leveraging anatomical features in fundus images that are relatively stable with respect to glaucomatous changes, the model accurately reconstructed individualized normative RNFLT maps, achieving high agreement with measured RNFLT (mean R^2^ = 0.81 ± 0.13). These results suggest that subject-specific normative RNFLT profiles can be reliably inferred from en face fundus anatomy, offering an alternative to conventional population-based normative references that do not account for inter-individual variability. A key motivation for our approach arises from the distinct temporal behaviors of retinal anatomy and RNFLT measurements. While RNFLT progressively changes with glaucomatous damage, the large-scale anatomical structures captured by en face fundus images, such as optic disc morphology and vessel configuration, remain largely stable within an individual. By training the model on eyes with normal visual fields, a mapping is learned from stable anatomy to personalized RNFLT norms. This enables prediction of the expected normal RNFLT from follow-up en face images, even as actual RNFLT declines, effectively separating anatomical baseline estimation from disease-related structural loss. The three examples (**Figure 7**) showed that the deep learning model can reasonably predict the normal RNFLT map from the corresponding OCT fundus image, even though raw RNFLT maps showed the retinal anatomy was severely damaged

**Figure 7.**
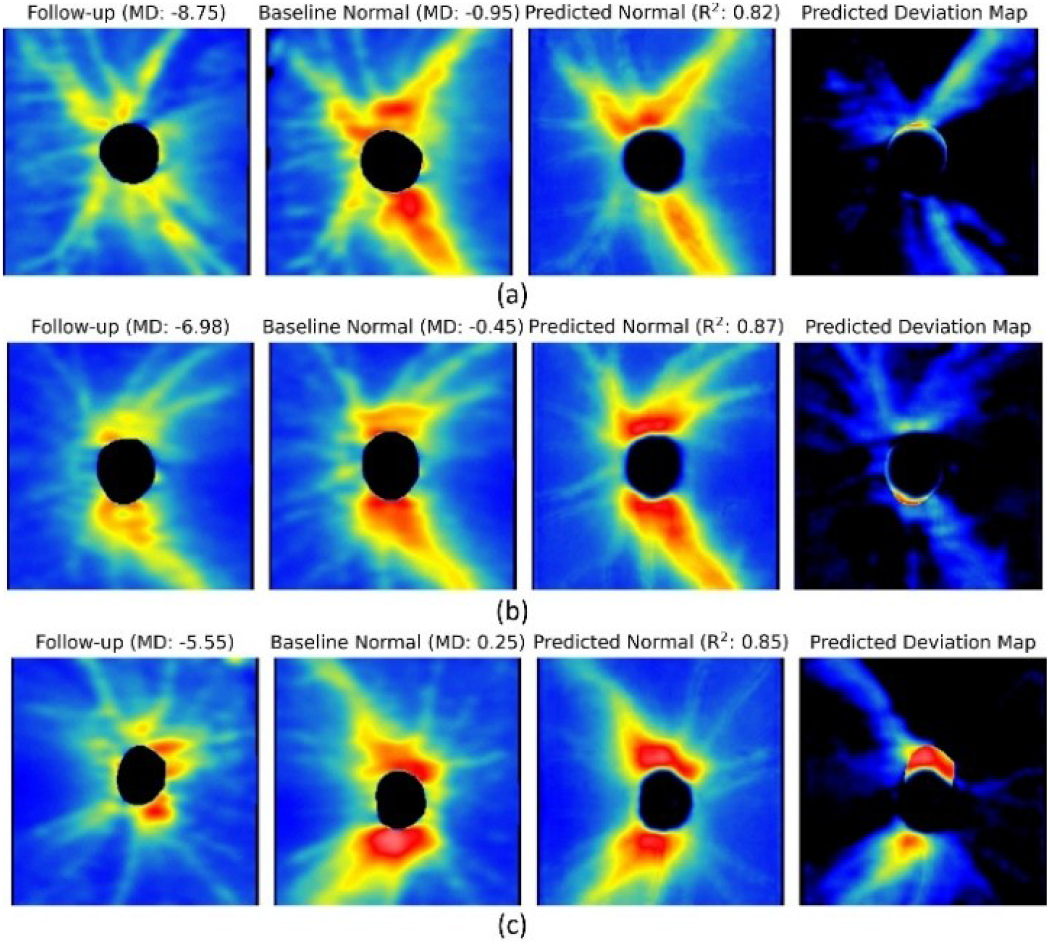
Representative examples showing predicted normative RNFLT maps and derived deviation maps in eyes with varying degrees of structural damage.

Using these individualized norms, RNFLT deviation maps demonstrated substantially stronger structure-function correlations than conventional RNFLT measurements. In linear regression analysis, the correlation between predicted deviation maps and VF MD reached R = 0.42, nearly doubling the correlation achieved with raw RNFLT maps (R = 0.19). This finding indicates that personalized RNFLT deviation maps more accurately reflect functional impairment and help explain the structure-function discrepancies commonly observed in clinical practice. The advantage of personalized RNFLT deviation representation was further confirmed in deep learning-based prediction of 52 visual field total deviation values. Models combining original RNFLT maps with RNFLT deviation maps achieved the lowest prediction error (mean MAE = 3.31 dB) and highest explanatory power (mean R^2^ = 0.39), outperforming models using either representation alone. This suggests that absolute RNFLT information and individualized deviation information provide complementary cues for functional prediction. Note that RNFLT deviation maps can predict VF TD maps with nearly the same accuracy as the original RNFLT maps (R^2^ 0.34 versus 0.35). The slight decrease of R^2^ may be due to slight anatomical changes due to glaucoma. For instance, previous research has reported that retinal blood vessels shift toward the nasal direction in glaucoma patients compared with normal subjects.^17^ Overall, our individualized RNFLT deviation maps can visualize patient-specific regions of thinning to assist glaucoma specialists.

Previous studies have attempted to establish more accurate RNFLT normative models by accounting for retinal anatomical variation, including sex, myopia, retinal blood vessel location, and optic disc parameters.^4, 7-9, 11, 18^ However, these approaches typically rely on manual measurements or selected anatomical features and are not designed to learn comprehensive, patient-specific normative RNFLT profiles.^9, 11, 19-21^ In parallel, machine learning and deep learning methods have been increasingly applied to glaucoma assessment, primarily for disease classification or functional prediction, yet most such models lack anatomically personalized normative modeling. ^22-24^ Consequently, existing AI-based frameworks either ignore individual anatomical variability or incorporate it only partially, limiting their ability to accurately characterize structure–function relationships. ^25-28^ The present study addresses this gap by directly learning patient-specific RNFLT norms from fundus anatomy using deep learning and leveraging derived deviation maps to improve structure–function correlation.

Clinically, classical age-based normative systems may mask glaucomatous damage in some eyes, particularly when RNFLT values fall within standard “normal” ranges. Personalized RNFLT maps revealed localized thinning relative to each individual’s expected anatomical profile, highlighting the risk of false-negative interpretations when anatomical variability is ignored. This approach may be especially valuable for early-stage glaucoma, atypical anatomy, and longitudinal disease monitoring.^29^

Several limitations should be acknowledged. The model was trained on eyes with normal visual fields, and its performance in other retinal or optic nerve diseases remains to be determined. In addition, this retrospective, single-center study may limit generalizability. Future studies incorporating multi-center data, broader disease spectra, and longitudinal follow-up are warranted.

## Conclusion

This study demonstrates that deep learning can accurately predict RNFLT norms from en face OCT fundus images. The predicted RNFLT norm and deviation maps show substantially improved structure-function correlation with VFs compared with conventional RNFLT measurements, particularly when combined with original RNFLT information. These findings suggest that deep learning-derived RNFLT norms can provide individualized visualization of RNFLT thinning regions with the potential to assist ophthalmologists in improving glaucoma diagnosis and monitoring in clinical practice.^29,30^

## Data Availability

The data are private, internal, and owned by Massachusetts Eye and Ear.

